# Evaluation of antithrombotic use and COVID-19 outcomes in a nationwide atrial fibrillation cohort

**DOI:** 10.1101/2021.09.03.21263023

**Authors:** Alex Handy, Amitava Banerjee, Angela Wood, Caroline E Dale, Cathie Sudlow, Christopher Tomlinson, Daniel Bean, Johan H Thygesen, Mehrdad A Mizani, Michail Katsoulis, Reecha Sofat, Richard Dobson, Rohan Takhar, Sam Hollings, Spiros Denaxas, Venexia Walker, on behalf of the CVD-COVID-UK Consortium

## Abstract

**Objective:** Evaluate antithrombotic (AT) use in individuals with atrial fibrillation (AF) and high stroke risk (CHA_2_DS_2_-VASc score>=2) and investigate whether pre-existing AT use may improve COVID-19 outcomes.

**Methods:** Individuals with AF and a CHA_2_DS_2_-VASc score>=2 on January 1^st^ 2020 were identified using pseudonymised, linked electronic health records for 56 million people in England and followed-up until May 1^st^ 2021. Factors associated with pre-existing AT use were analysed using logistic regression. Differences in COVID-19 related hospitalisation and death were analysed using logistic and Cox regression for individuals exposed to pre-existing AT use vs no AT use, anticoagulants (AC) vs antiplatelets (AP) and direct oral anticoagulants (DOACs) vs warfarin.

**Results:** From 972,971 individuals with AF and a CHA_2_DS_2_-VASc score>=2, 88.0% (n=856,336) had pre-existing AT use, 3.8% (n=37,418) had a COVID-19 related hospitalisation and 2.2% (n=21,116) died. Factors associated with no AT use included comorbidities that may contraindicate AT use (liver disease and history of falls) and demographics (socioeconomic status and ethnicity). Pre-existing AT use was associated with lower odds of death (OR=0.92 *[0*.*87-0*.*96 at 95% CI]*), but higher odds of hospitalisation OR=1.20 *[1*.*15-1*.*26 at 95% CI]*). The same pattern was observed for AC vs AP (death (OR=0.93 [0.87-0.98]), hospitalisation (OR=1.17 [1.11-1.24])) but not for DOACs vs warfarin (death (OR=1.00 [0.95-1.05]), hospitalisation (OR=0.86 [0.82-0.89]).

**Conclusions:** Pre-existing AT use may offer marginal protection against COVID-19 death, with AC offering more protection than AP. Although this association may not be causal, it provides further incentive to improve AT coverage for eligible individuals with AF.

**KEY QUESTIONS:** *What is already known about this subject?:*

- Anticoagulants (AC), a sub-class of antithrombotics (AT), reduce the risk of stroke and are recommended for individuals with atrial fibrillation (AF) and at high risk of stroke (CHA_2_DS_2_-VASc score>=2, National Institute for Health and Care Excellence threshold). However, previous evaluations suggest that up to one third of these individuals may not be taking AC. Over estimation of bleeding and fall risk in elderly patients have been identified as potential factors in this under medicating.
- In response to the COVID-19 pandemic, several observational studies have observed correlations between pre-existing AT use, particularly anticoagulants (AC), and lower risk of severe COVID-19 outcomes such as hospitalisation and death. However, these correlations are inconsistent across studies and have not compared all major sub-types of AT in one study.

*What does this study add?:*

- This study uses datasets covering primary care, secondary care, pharmacy dispensing, death registrations, multiple COVID-19 diagnoses routes and vaccination records for 56 million people in England and is the largest scale evaluation of AT use to date. This provides the statistical power to robustly analyse targeted sub-types of AT and control for a wide range of potential confounders. All code developed for the study is opensource and an updated nationwide evaluation can be rapidly created for future time points.
- In 972,971 individuals with AF and a CHA_2_DS_2_-VASc score>=2, we observed 88.0% (n=856,336) with pre-existing AT use which was associated with marginal protection against COVID-19 death (OR=0.92 *[0*.*87-0*.*96 at 95% CI]*).

*How might this impact on clinical practice?:*

- These findings can help shape global AT medication policy and provide population-scale, observational analysis results alongside gold-standard randomised control trials to help assess whether a potential beneficial effect of pre-existing AT use on COVID-19 death alters risk to benefit assessments in AT prescribing decisions.

## INTRODUCTION

Atrial fibrillation (AF) is a disturbance of heart rhythm affecting 37.5 million people globally[1] and significantly increases stroke risk[2]. Anticoagulants (AC), a sub-type of antithrombotics (AT), reduce the risk of stroke[3] and are recommended for individuals with AF and a high risk of stroke (CHA_2_DS_2_-VASc score>=2, National Institute for Health and Care Excellence (NICE) threshold)[4,5]. Despite improvements in AC uptake, previous evaluations suggest that up to one third of individuals with AF and a CHA_2_DS_2_-VASc score>=2 in the UK may not be on AC[6], with around 15% on no type of AT[6]. Hypotheses for this sub-optimal medication centre around clinical over estimation of bleeding and fall risk in elderly patients[6,7] but the potential drivers of AT use remain under explored at population scale.

COVID-19 has presented another risk factor for individuals with AF, who are at increased risk of poor outcomes if they become infected[8]. Observational evidence from Germany (n=6,637) suggests that pre-existing AC use, but not antiplatelets (AP - another sub-type of AT), may reduce mortality for individuals hospitalised with COVID-19[9]. However, evidence is discordant with a US study (n=3,772) observing no difference in mortality in groups on AC or AP[10]. In the UK, a larger study (n = 70,464 / 372,746) explored AC and AC sub-types (warfarin vs direct oral anticoagulants (DOACs)) in individuals with AF and observed that AC was associated with a lower COVID-19 specific mortality[11]. This observational evidence is promising, but it does not compare all sub-types of AT and only covered the period up to September 28^th^ 2020.

This study, therefore, set out to conduct the largest scale evaluation of AT use in individuals with AF to date in routinely updated, linked, population-scale electronic health record (EHR) data for 56 million people in England[12]. Utilising this statistical power, this study investigated what factors are associated with pre-existing AT use and whether pre-existing AT use (across sub-types) may reduce COVID-19 related hospitalisation and death.

## METHODS

### Study design and data sources

We conducted a cohort analysis using the newly established NHS Digital Trusted Research Environment (TRE) for England which provides secure, remote access to linked, person level EHR data for over 56 million people[12]. Available data sources cover primary care, secondary care, pharmacy dispensing, death registrations and COVID-19 tests and vaccines. For this study, we used the General Practice Extraction Service Extract for Pandemic Planning and Research (GDPPR) for demographic and diagnostic data (e.g. a diagnosis of AF) and the NHS Business Service Authority Dispensed Medicines (BSADM) for medication exposure data (e.g. pre-existing AT use) as this is the most accurate available representation of the medication an individual takes. Hospital Episode Statistics (HES), COVID-19 Hospitalisations in England Surveillance System (CHESS), Secondary Uses Service (SUS) and Office for National Statistics (ONS) Civil Registration of Deaths were used for COVID-19 hospitalisation and death. Public Health England’s Second Generation Surveillance System (SGSS) was used to identify COVID-19 test results and the COVID-19 vaccination events dataset was used for COVID-19 vaccine status.

*Figure 1* provides an illustrative overview of how cohorts were designed to address the study objectives and the open protocol (https://github.com/BHFDSC/CCU020/tree/main/england/protocol) provides an overview, including all protocol amendments.

**Figure 1:**
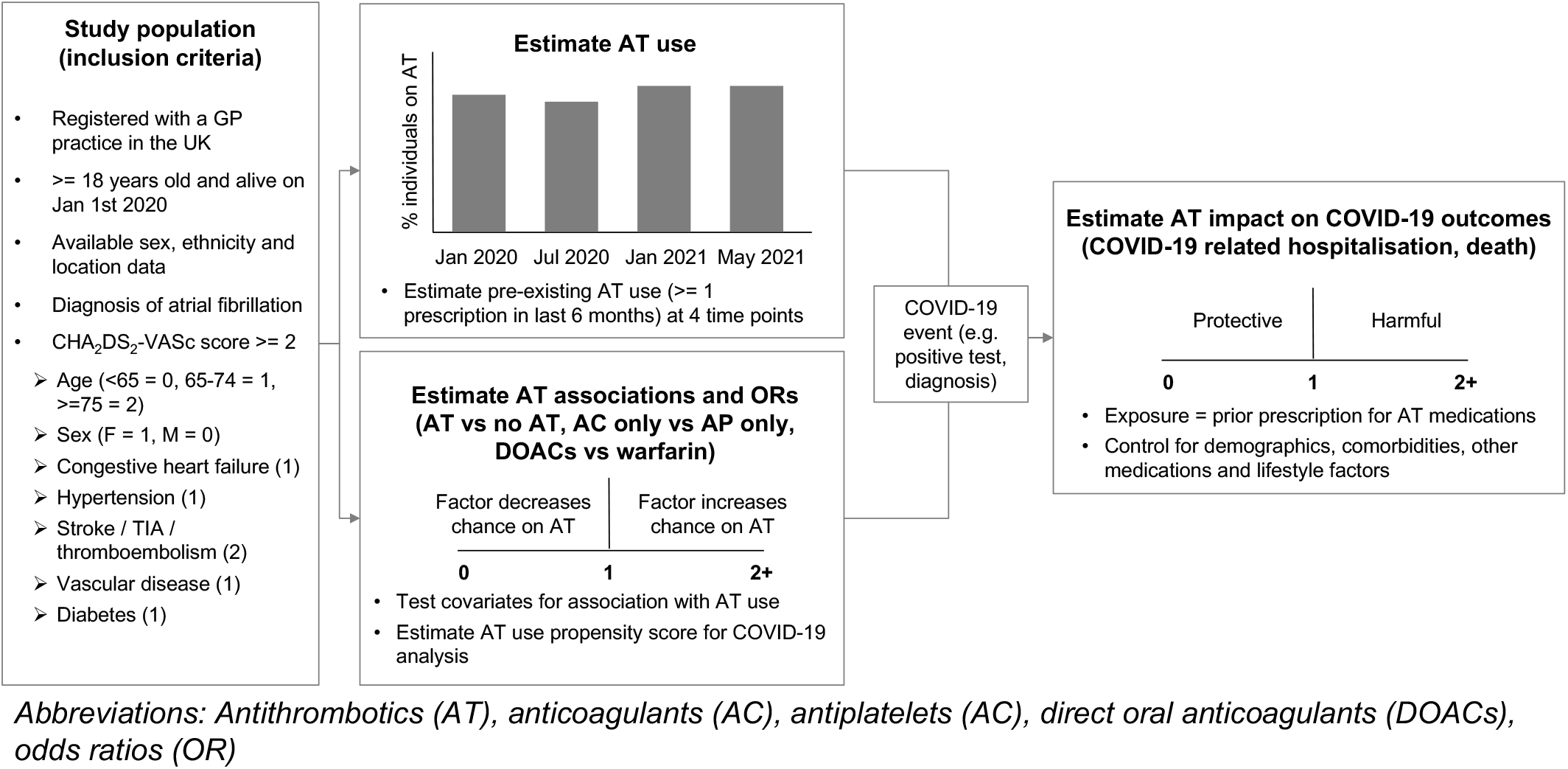
Illustrative overview of study design.

### Study populations

Individuals were included in the study if registered with a GP practice in England (at least one record in the GDPPR dataset with a valid person pseudo identifier), >= 18 years old and alive on January 1st 2020, had available sex, ethnicity and GP practice location data (based on most recent, available data across primary care (GDPPR), secondary care (HES) and death registrations (ONS)) and had a diagnosis of AF (coded in GDPPR) with a CHA_2_DS_2_-VASc score>=2 (calculated from the sum of components[13] coded in GDPPR).

Individuals with contraindications for sub-types of AT (e.g. DOACs in mitral stenosis, prosthetic mechanical valves, antiphospholipid antibody syndrome) were included as they are still eligible for other AT sub-types (e.g. AP, warfarin).

To investigate exposure to pre-existing AT use on COVID-19 related hospitalisation and death, inclusion criteria of a recorded COVID-19 event were applied. A COVID-19 event was defined as any of a positive test (polymerase chain reaction or lateral flow), a coded diagnosis in primary or secondary care or a COVID-19 diagnosis on a death certificate (see Thygesen et al (https://github.com/BHFDSC/CCU013_01_ENG-COVID-19_event_phenotyping/tree/main/phenotypes) for further details and phenotyping algorithms).

All phenotyping algorithms used are available on Github (https://github.com/BHFDSC/CCU020/tree/main/england/phenotypes). *Supplementary Figure 1* provides an overview of the numbers of individuals excluded at each stage.

### Study variables

#### Medication exposure

An individual was defined as taking a particular medication if they had >=1 dispensed prescription (coded in the NHS BSADM) in the previous 6 months. We purposefully defined a liberal threshold to support evaluation of AT usage up to May 2021 that may have included unusual buying patterns (e.g. bulk buying) caused by the pandemic.

Mutually exclusive medication categories were constructed for AC only, AP only, AP and AC and no AT. Apixaban, rivaroxaban, dabigatran and edoxaban were collectively categorised as DOACs for comparison with warfarin. For analysis, three mutually exclusive medication categories were tested (any AT vs no AT, AC only vs AP only, DOACs vs warfarin).

#### Outcomes

We defined two COVID-19 outcomes, COVID-19 related hospitalisation and COVID-19 death. COVID-19 hospitalisation included any hospital admission with a recorded COVID-19 diagnosis in any position (e.g. not the primary diagnosis). COVID-19 death included individuals with a COVID-19 diagnosis on their death certificate in any position, a registered death within 28 days of their first recorded COVID-19 event or a discharge destination denoting death after a COVID-19 hospitalisation (see Thygesen et al (https://github.com/BHFDSC/CCU013_01_ENG-COVID-19_event_phenotyping/tree/main/phenotypes) for further details and phenotyping algorithms). Follow-up for COVID-19 outcomes ended on 1^st^ May 2021 with the final follow-up date as either the date of the outcome of interest (e.g. COVID-19 death) or the study end date (1^st^ May 2021).

#### Covariates

Covariates were pre-selected based on potential associations with pre-existing AT use[6] or COVID-19 outcomes and included demographics *(age, sex, ethnicity, geographic location, socio-economic status (as measured by index of multiple deprivation (IMD) decile))*, comorbidities that increase stroke and bleeding risk *(congestive heart failure, hypertension, stroke, vascular disease, diabetes, uncontrolled hypertension, renal disease, liver disease, prior major bleeding, hazardous alcohol use, history of fall, body mass index (BMI), smoking status)* and other medications *(anti-hypertensives, lipid-regulating drugs, proton pump inhibitors, NSAIDs, corticosteroids, other immunosuppressants and COVID-19 vaccination status, defined as at least one vaccine recorded in the COVID-19 vaccination events dataset prior to the individual’s COVID-19 event)*.

The same covariates (excluding COVID-19 vaccination status) were used as independent variables to test associations with pre-existing AT use (for any AT vs no AT, AP only vs AC only, DOACs vs warfarin) and to calculate a propensity score for use as an additional covariate in the COVID-19 outcome analysis.

### Statistical analysis

Descriptive statistics were used to summarise the study population characteristics and were stratified by medication category. Pairwise Pearson’s correlation coefficients were used to check for potential collinearities between covariates. Multivariable logistic regression was used to test associations with pre-existing AT use and calculate the propensity score.

Multivariable logistic regression and Cox regression were used to test differences between exposure groups (any AT vs no AT, AC only vs AP only, DOACs vs warfarin) for COVID-19 related hospitalisation and death. These methods were selected to evaluate potential differences between event (logistic regression) and time-to-event (Cox regression) based analysis. All covariates including the propensity score were included in both methods. For variables with incomplete data (BMI – 9.3% missing), individual values were imputed with the cohort mean.

Two sensitivity analyses were conducted. Firstly, to evaluate the potential impact of different time periods, analysis was repeated for Jan 1^st^ 2020 – December 1^st^ 2020, prior to the introduction of vaccines and the December 29^th^ 2020 cases peak of the second wave[14]. Secondly, to validate the potential effect on COVID-19 specific outcomes, analysis was repeated with COVID-19 hospitalisation and death defined exclusively as the primary recorded diagnosis (coded first in hospital record or death certificate).

Primary results are reported from the multivariable logistic regression models covering the full time period (Jan 1^st^ 2020 – May 1^st^ 2021) with the other analyses reviewed for concordance.

Data preparation was performed using Python 3.7 and Spark SQL (2.4.5) on Databricks Runtime 6.4 for Machine Learning with analysis performed using R version 4.0.3. All code for data preparation and analysis is available on Github (https://github.com/BHFDSC/CCU020/tree/main/england/code) and full results available at this microsite (https://alexhandy1.shinyapps.io/at-evaluation-results/)

### Ethical and Regulatory Approvals

Data access approval was granted to the CVD-COVID-UK consortium through the NHS Digital online Data Access Request Service (ref. DARS-NIC-381078-Y9C5K). The BHF Data Science Centre approvals and oversight board deemed that this project (proposal CCU020 Evaluation of antithrombotic use and COVID-19 outcomes) fell within the scope of the consortium’s ethical and regulatory approvals. Analyses were conducted by approved researcher (AH) via secure remote access to the TRE. Only summarised, aggregate results were exported, following manual review by the NHS Digital ‘safe outputs’ escrow service, to ensure no output placed in the public domain contains information that may be used to identify an individual[12]. The North East-Newcastle and North Tyneside 2 research ethics committee provided ethical approval for the CVD-COVID-UK research programme (REC No 20/NE/0161).

## RESULTS

### Evaluation of antithrombotic use

From a total of 55,903,113 individuals registered with a GP practice in England, 972,971 (1.7%) had a diagnosis of AF and a CHA_2_DS_2_-VASc score>=2 on January 1^st^ 2020 and 88.0% (n=856,336) had pre-existing AT use, with 74.3% (n=722,737) on AC only. The demographic and clinical characteristics of this cohort are summarised in *Tables 1-3*. By May 2021, the proportion of individuals on any AT had fallen to 87.7% but AC only had increased to 75.7% (see *Figure 2*). For individuals on any AT, warfarin prescriptions fell from 24.8% in January 2020 to 17.1% in May 2021 whilst DOACs rose from 60.3% to 69.5% (see *Supplementary Figure 2*).

**Table 1.** study population demographic characteristics by antithrombotic medication category. Percentages should be interpreted vertically for all variables e.g. proportion within category for variable, except for the first row showing percentage of individuals across AT medication categories. Any AT = any antithrombotic, AC only = anticoagulants only, AP only = antiplatelets only, AC and AP = anticoagulants and antiplatelets, no AT = no antithrombotic.

|  | Total | Any AT | AC only | AP only | AC and AP | No AT |
| --- | --- | --- | --- | --- | --- | --- |
| Individuals | 972971<br>(100%) | 856336<br>(88%) | 722737<br>(74.3%) | 70498<br>(7.2%) | 63101<br>(6.5%) | 116635<br>(12%) |
| Age (mean years, +/- sd) | 79 (+/- 9.3) | 79 (+/- 9) | 79 (+/- 8.9) | 79 (+/- 10) | 78 (+/- 8.9) | 78 (+/- 11) |
| 65-74 | 229464<br>(23.6%) | 198956<br>(23.2%) | 166943<br>(23.1%) | 16018<br>(22.7%) | 15995<br>(25.3%) | 30508<br>(26.2%) |
| $\geq 75$ | 686578<br>(70.6%) | 610497<br>(71.3%) | 518205<br>(71.7%) | 49702<br>(70.5%) | 42590<br>(67.5%) | 76081<br>(65.2%) |
| Female | 449279<br>(46.2%) | 387184<br>(45.2%) | 338477<br>(46.8%) | 28622<br>(40.6%) | 20085<br>(31.8%) | 62095<br>(53.2%) |
| Ethnicity |  |  |  |  |  |  |
| White | 932571<br>(95.8%) | 822292<br>(96%) | 696757<br>(96.4%) | 66237<br>(94%) | 59298<br>(94%) | 110279<br>(94.6%) |
| Asian or Asian British | 20557<br>(2.1%) | 17699<br>(2.1%) | 12797<br>(1.8%) | 2536<br>(3.6%) | 2366<br>(3.7%) | 2858<br>(2.5%) |
| Black or Black British | 9418<br>(1%) | 7658<br>(0.9%) | 6200<br>(0.9%) | 862<br>(1.2%) | 596<br>(0.9%) | 1760<br>(1.5%) |
| Mixed | 3194<br>(0.3%) | 2636<br>(0.3%) | 2115<br>(0.3%) | 274<br>(0.4%) | 247<br>(0.4%) | 558<br>(0.5%) |
| Other Ethnic Groups | 7231<br>(0.7%) | 6051<br>(0.7%) | 4868<br>(0.7%) | 589<br>(0.8%) | 594<br>(0.9%) | 1180<br>(1%) |
| Geographical locations |  |  |  |  |  |  |
| South East | 172714<br>(17.8%) | 150276<br>(17.5%) | 127207<br>(17.6%) | 11566<br>(16.4%) | 11503<br>(18.2%) | 22438<br>(19.2%) |
| North West | 143391<br>(14.7%) | 127860<br>(14.9%) | 106990<br>(14.8%) | 10705<br>(15.2%) | 10165<br>(16.1%) | 15531<br>(13.3%) |
| East of England | 104591<br>(10.7%) | 92676<br>(10.8%) | 78194<br>(10.8%) | 7408<br>(10.5%) | 7074<br>(11.2%) | 11915<br>(10.2%) |
| South West | 108250<br>(11.1%) | 94816<br>(11.1%) | 80009<br>(11.1%) | 7863<br>(11.2%) | 6944<br>(11%) | 13434<br>(11.5%) |
| Yorkshire and The<br>Humber | 108285<br>(11.1%) | 96113<br>(11.2%) | 81386<br>(11.3%) | 8405<br>(11.9%) | 6322<br>(10%) | 12172<br>(10.4%) |
| West Midlands | 111062<br>(11.4%) | 97555<br>(11.4%) | 83383<br>(11.5%) | 7836<br>(11.1%) | 6336<br>(10%) | 13507<br>(11.6%) |
| East Midlands | 83786<br>(8.6%) | 74596<br>(8.7%) | 63978<br>(8.9%) | 5773<br>(8.2%) | 4845<br>(7.7%) | 9190<br>(7.9%) |
| London | 95746<br>(9.8%) | 81824<br>(9.6%) | 66815<br>(9.2%) | 7495<br>(10.6%) | 7514<br>(11.9%) | 13922<br>(11.9%) |
| North East | 45146<br>(4.6%) | 40620<br>(4.7%) | 34775<br>(4.8%) | 3447<br>(4.9%) | 2398<br>(3.8%) | 4526<br>(3.9%) |
| IMD deciles |  |  |  |  |  |  |
| 1 (most deprived) | 78061<br>(8%) | 68894<br>(8%) | 56583<br>(7.8%) | 6490<br>(9.2%) | 5821<br>(9.2%) | 9167<br>(7.9%) |
| 10 (least deprived) | 106436<br>(10.9%) | 93984<br>(11%) | 80764<br>(11.2%) | 6795<br>(9.6%) | 6425<br>(10.2%) | 12452<br>(10.7%) |

**Table 2.** study population comorbidities that increase stroke and bleeding risk by antithrombotic medication category. Percentages should be interpreted vertically for all variables e.g. proportion within category for variable. HAS-BLED score component bleeding medications excluded as measured within exposures and labile INR excluded as could not be accurately extracted from datasets.

|  | Total | Any AT | AC only | AP only | AC and AP | No AT |
| --- | --- | --- | --- | --- | --- | --- |
| CHA2DS2-VASc score components |  |  |  |  |  |  |
| Vascular disease | 169797<br>(17.5%) | 159892<br>(18.7%) | 103946<br>(14.4%) | 23815<br>(33.8%) | 32131<br>(50.9%) | 9905<br>(8.5%) |
| Stroke / TIA / Thromboembolism | 196899<br>(20.2%) | 183140<br>(21.4%) | 150588<br>(20.8%) | 16611<br>(23.6%) | 15941<br>(25.3%) | 13759<br>(11.8%) |
| Congestive heart failure | 247562<br>(25.4%) | 228877<br>(26.7%) | 192023<br>(26.6%) | 15038<br>(21.3%) | 21816<br>(34.6%) | 18685<br>(16%) |
| Diabetes | 268437<br>(27.6%) | 242060<br>(28.3%) | 197216<br>(27.3%) | 21602<br>(30.6%) | 23242<br>(36.8%) | 26377<br>(22.6%) |
| Hypertension | 675676<br>(69.4%) | 600623<br>(70.1%) | 505514<br>(69.9%) | 49678<br>(70.5%) | 45431<br>(72%) | 75053<br>(64.3%) |
| CHA2DS2-VASc score (mean, +/- sd) | 3.9 (+/- 1.4) | 4 (+/- 1.4) | 3.9 (+/- 1.4) | 4.1 (+/- 1.5) | 4.4 (+/- 1.5) | 3.4 (+/- 1.3) |
| 2 | 172172<br>(17.7%) | 138750<br>(16.2%) | 120969<br>(16.7%) | 10912<br>(15.5%) | 6869<br>(10.9%) | 33422<br>(28.7%) |
| 3 | 245979<br>(25.3%) | 213057<br>(24.9%) | 184241<br>(25.5%) | 16289<br>(23.1%) | 12527<br>(19.9%) | 32922<br>(28.2%) |
| 4 | 252047<br>(25.9%) | 224255<br>(26.2%) | 190707<br>(26.4%) | 17875<br>(25.4%) | 15673<br>(24.8%) | 27792<br>(23.8%) |
| 5 | 162318<br>(16.7%) | 149105<br>(17.4%) | 122356<br>(16.9%) | 12995<br>(18.4%) | 13754<br>(21.8%) | 13213<br>(11.3%) |
| >=6 | 140455<br>(14.4%) | 131169<br>(15.3%) | 104464<br>(14.5%) | 12427<br>(17.6%) | 14278<br>(22.6%) | 9286<br>(8%) |
| HAS-BLED score components |  |  |  |  |  |  |
| Renal disease | 315940<br>(32.5%) | 284379<br>(33.2%) | 237965<br>(32.9%) | 24423<br>(34.6%) | 21991<br>(34.9%) | 31561<br>(27.1%) |
| Liver disease | 8462<br>(0.9%) | 6707<br>(0.8%) | 5440<br>(0.8%) | 788<br>(1.1%) | 479<br>(0.8%) | 1755<br>(1.5%) |
| Stroke | 196493<br>(20.2%) | 182756<br>(21.3%) | 150232<br>(20.8%) | 16606<br>(23.6%) | 15918<br>(25.2%) | 13737<br>(11.8%) |
| Major bleeding event | 335289<br>(34.5%) | 293096<br>(34.2%) | 240703<br>(33.3%) | 27431<br>(38.9%) | 24962<br>(39.6%) | 42193<br>(36.2%) |
| Harmful alcohol use | 28970<br>(3%) | 25572<br>(3%) | 21162<br>(2.9%) | 2274<br>(3.2%) | 2136<br>(3.4%) | 3398<br>(2.9%) |
| Uncontrolled hypertension | 66576<br>(6.8%) | 58873<br>(6.9%) | 48444<br>(6.7%) | 5395<br>(7.7%) | 5034<br>(8%) | 7703<br>(6.6%) |
| History of fall | 119738<br>(12.3%) | 103615<br>(12.1%) | 85718<br>(11.9%) | 10717<br>(15.2%) | 7180<br>(11.4%) | 16123<br>(13.8%) |
| BMI (mean, +/- sd) | 28.7 (+/- 6) | 28.8 (+/- 6) | 28.8 (+/- 6.1) | 28.1 (+/- 5.6) | 29 (+/- 5.8) | 27.9 (+/- 5.9) |
| Smoking status (ever smoker) | 638775<br>(65.7%) | 566860<br>(66.2%) | 472208<br>(65.3%) | 48567<br>(68.9%) | 46085<br>(73%) | 71915<br>(61.7%) |

**Table 3.** study population characteristics for COVID-19 outcomes and other medications by antithrombotic medication category. Percentages should be interpreted vertically for all variables e.g. proportion within category for variable. Pre-existing medication use was determined as >=1 dispensed prescription in the 6 months prior to the cohort start date (Jan 1st 2020).

|  | Total | Any AT | AC only | AP only | AC and AP | No AT |
| --- | --- | --- | --- | --- | --- | --- |
| COVID-19 outcomes |  |  |  |  |  |  |
| Any COVID-19 event | 77364<br>(8%) | 67087<br>(7.8%) | 54756<br>(7.6%) | 6743<br>(9.6%) | 5588<br>(8.9%) | 10277<br>(8.8%) |
| COVID-19 hospitalisation | 37418<br>(3.8%) | 33150<br>(3.9%) | 26887<br>(3.7%) | 3201<br>(4.5%) | 3062<br>(4.9%) | 4268<br>(3.7%) |
| COVID-19 hospitalisation (primary diagnosis) | 27011<br>(2.8%) | 23919<br>(2.8%) | 19375<br>(2.7%) | 2319<br>(3.3%) | 2225<br>(3.5%) | 3092<br>(2.7%) |
| COVID-19 death | 21116<br>(2.2%) | 18173<br>(2.1%) | 14553<br>(2%) | 2055<br>(2.9%) | 1565<br>(2.5%) | 2943<br>(2.5%) |
| COVID-19 death<br>(primary diagnosis) | 15297<br>(1.6%) | 13158<br>(1.5%) | 10522<br>(1.5%) | 1508<br>(2.1%) | 1128<br>(1.8%) | 2139<br>(1.8%) |
| Other medications |  |  |  |  |  |  |
| Antihypertensives | 540678<br>(55.6%) | 498113<br>(58.2%) | 412077<br>(57%) | 40375<br>(57.3%) | 45661<br>(72.4%) | 42565<br>(36.5%) |
| Lipid regulating drugs | 589568<br>(60.6%) | 547521<br>(63.9%) | 441736<br>(61.1%) | 51120<br>(72.5%) | 54665<br>(86.6%) | 42047<br>(36.1%) |
| Proton pump inhibitors | 409429<br>(42.1%) | 369461<br>(43.1%) | 286984<br>(39.7%) | 39180<br>(55.6%) | 43297<br>(68.6%) | 39968<br>(34.3%) |
| NSAIDS | 19448<br>(2%) | 14608<br>(1.7%) | 11101<br>(1.5%) | 2317<br>(3.3%) | 1190<br>(1.9%) | 4840<br>(4.1%) |
| Corticosteroids | 80347<br>(8.3%) | 71706<br>(8.4%) | 59511<br>(8.2%) | 5929<br>(8.4%) | 6266<br>(9.9%) | 8641<br>(7.4%) |
| Other immunosuppressants | 13216<br>(1.4%) | 11690<br>(1.4%) | 9498<br>(1.3%) | 1152<br>(1.6%) | 1040<br>(1.6%) | 1526<br>(1.3%) |
| COVID-19 vaccine prior<br>to COVID-19 event | 9463<br>(1%) | 8248<br>(1%) | 6799<br>(0.9%) | 824<br>(1.2%) | 625 (1%) | 1215<br>(1%) |

**Figure 2:**
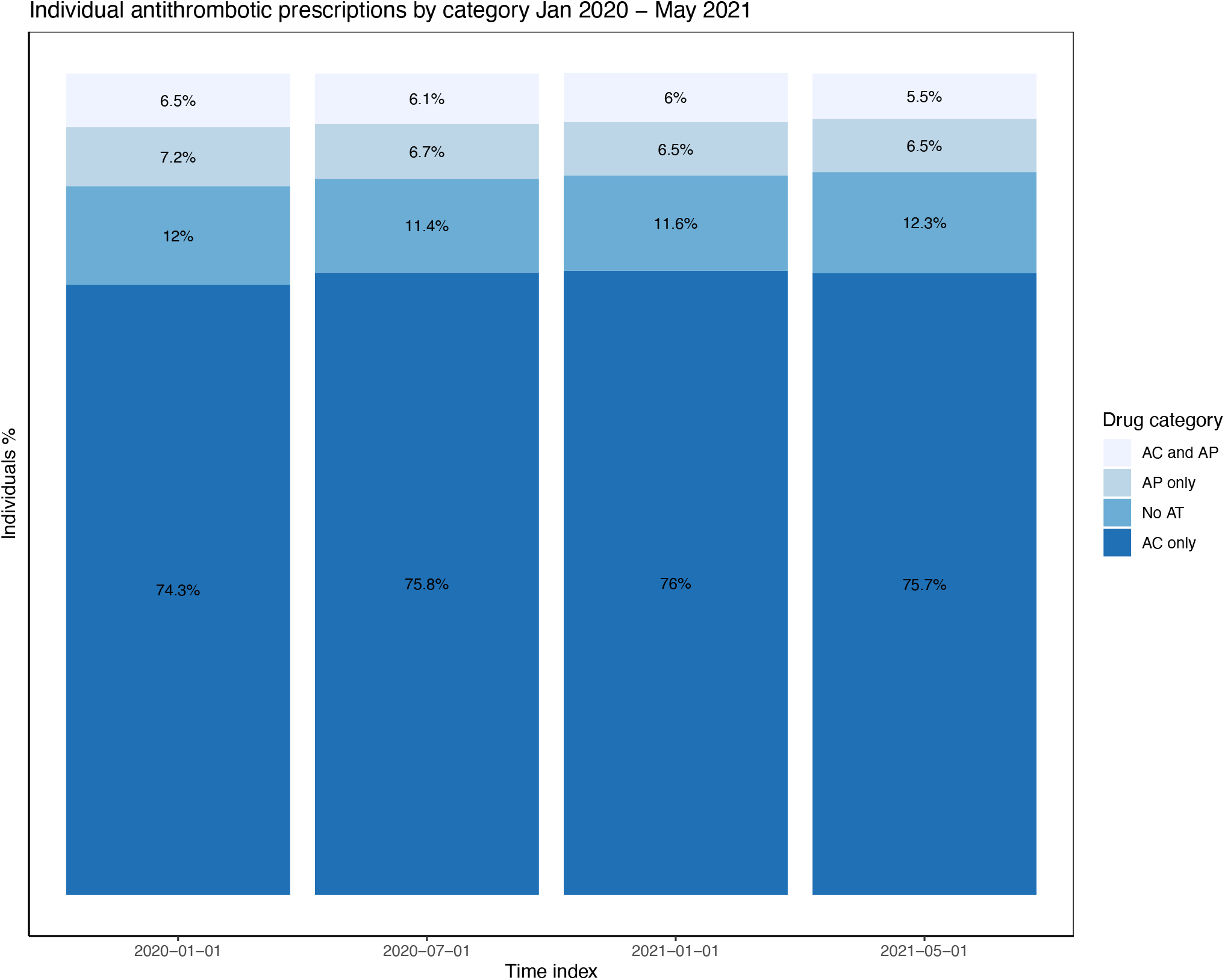
Individual antithrombotic prescriptions by category January 2020 – May 2021. Ordered by proportion of prescriptions with mutually exclusive categories.

Factors associated with pre-existing AT use vs no AT are shown in *Figure 3*. Lipid regulating drugs (OR=2.50 *[2*.*47-2*.*54 at 95% CI]*) and antihypertensives (OR=1.90 *[1*.*88-1*.*93 at 95% CI]*) were associated with the highest odds of pre-existing AT use followed by comorbidities in the CHA_2_DS_2_-VASc score (stroke (OR=1.76 *[1*.*72-1*.*79 at 95% CI]*, vascular disease (OR=1.60 *[1*.*56-1*.*63 at 95% CI])*. In contrast, nonsteroidal anti-Inflammatory drugs (NSAIDS, OR=0.39 *[0*.*37-0*.*40])*, liver disease (OR=0.56 *[0*.*53-0*.*59])* and history of falls (OR=0.80 *[0*.*79-0*.*82])* were associated with reduced odds.

**Figure 3:**
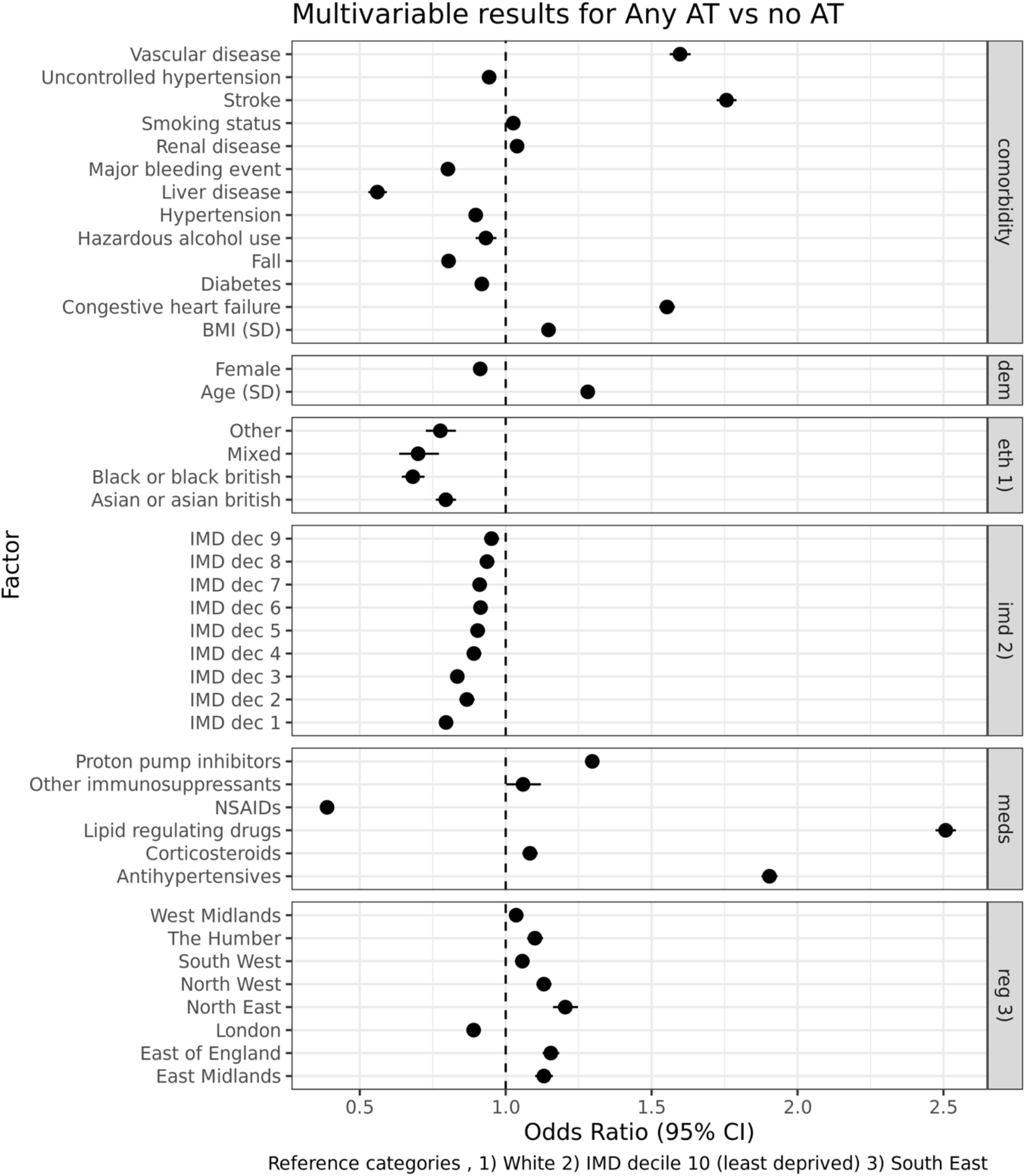
Factors associated with AT vs no AT medication (January 1^st^ 2020) using multivariable logistic regression.

Differences were also observed across demographics, ethnicity, socio-economic status and geographic location, with females (OR=0.91 *[0*.*90-0*.*92]*) and individuals from ethnic minorities and lower socio-economic positions associated with lower odds of AT use (e.g. ethnicity of black or black british vs white (OR=0.68 *[0*.*64-0*.*72]*). In other AT sub-types (AC vs AP and DOACs vs warfarin) results were broadly consistent (see *Supplementary Material Figures 3-4*), with the primary exception that vascular disease was associated with reduced odds of AC vs AP (OR=0.37 *[0*.*36-0*.*38*)]).

**Figure 4:**
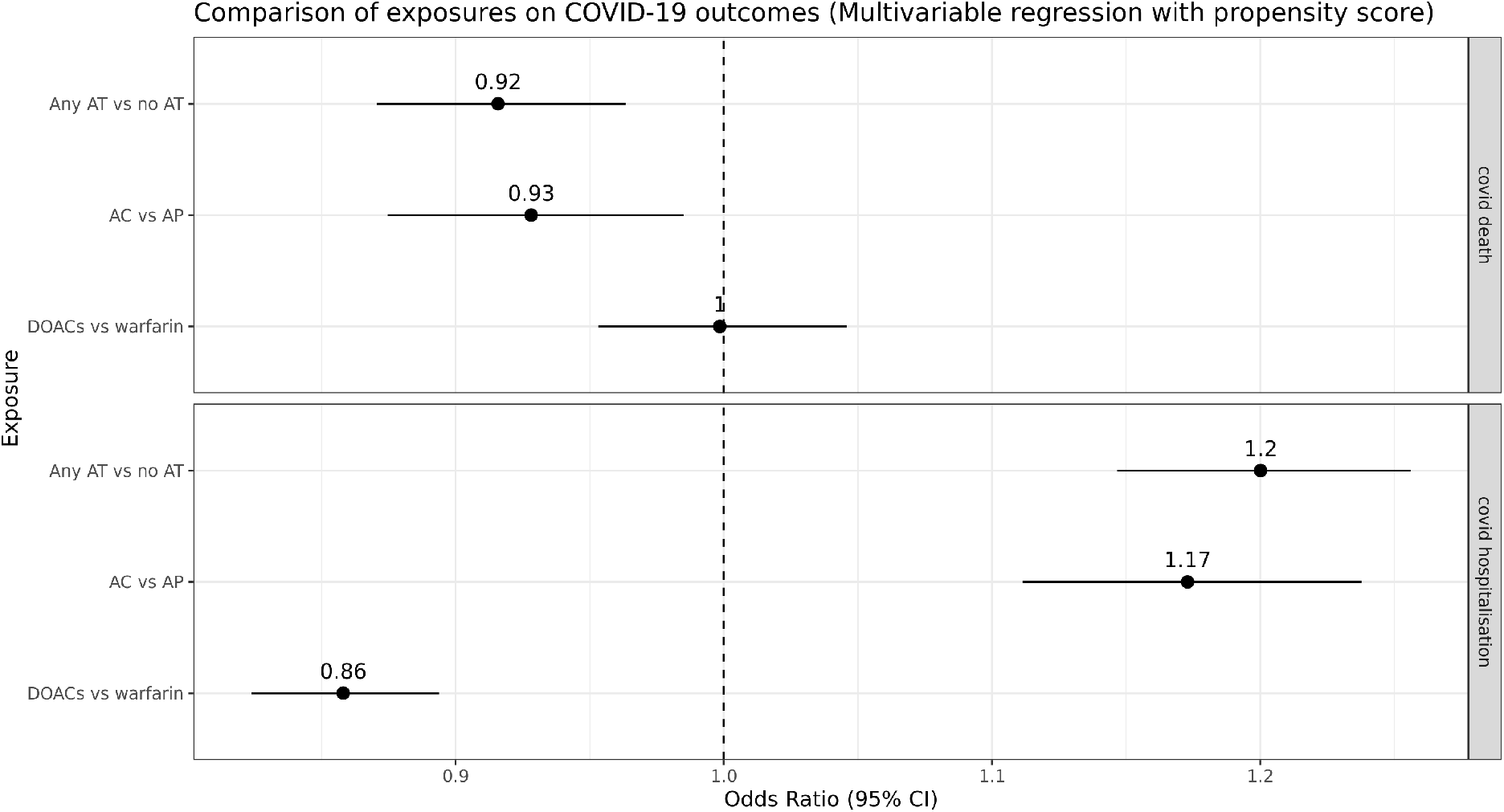
Comparison of AT medication exposures on COVID-19 outcomes (follow up to May 1^st^ 2021) using propensity score adjusted multivariable logistic regression.

### Antithrombotic use and COVID-19 outcomes

From 972,971 individuals that had a diagnosis of AF and a CHA_2_DS_2_-VASc score>=2 on January 1^st^ 2020, 8% (n=77,364) had a recorded COVID-19 event, 3.8% (n=37,418) had a COVID-19 related hospitalisation and 2.2% (n=21,116) died when followed up to 1^st^ May 2021. The summary characteristics of individuals with a recorded COVID-19 event are summarised in *Supplementary Tables 1-3*. Mean age (81) and comorbidities (mean CHA_2_DS_2_-VASc score 4.2) were both marginally higher compared to the full cohort. The proportion of individuals with pre-existing AT use was also marginally lower at 86.7%, but otherwise demographic and clinical characteristics were consistent.

Pre-existing AT use was associated with lower odds of COVID-19 death (OR=0.92 *[0*.*87-0*.*96 at 95% CI]*), but higher odds of COVID-19 hospitalisation OR=1.20 *[1*.*15-1*.*26 at 95% CI]*) (see *Figure 4*). The same pattern was observed for AC vs AP (COVID-19 death (OR=0.93 [0.87-0.98]), COVID-19 hospitalisation (OR=1.17 [1.11-1.24])) but not for DOACs vs warfarin (COVID-19 death (OR=1.00 [0.95-1.05]), COVID-19 hospitalisation (OR=0.86 [0.82-0.89]).

These results were all directionally consistent across Cox regression analysis and the sensitivity analyses (see *Supplementary Material Figure* 5-7). Full results are available on this microsite (https://alexhandy1.shinyapps.io/at-evaluation-results/).

## DISCUSSION

### Main findings

This study of routinely collected healthcare data from 56 million individuals in England is the largest scale evaluation of AT use to date. In 972,971 individuals with AF and a CHA_2_DS_2_-VASc score>=2, we observed 88.0% (n=856,336) with pre-existing AT use which was associated with marginal protection against COVID-19 death (OR=0.92 *[0*.*87-0*.*96 at 95% CI]*). Although this association may not be causal, it provides further incentive to improve AT coverage for eligible individuals with AF.

Of the AF cohort analysed, 8% (n=77,364) had a recorded COVID-19 event of which 3.8% (n=37,418) had a COVID-19 related hospitalisation and 2.2% (n=21,116) died. A marginally lower risk of COVID-19 death was observed for those with pre-existing AT use which directionally aligns with the most comparable previous studies[9,11]. AT use was, however, associated with higher odds of COVID-19 hospitalisation. This observation remained consistent when including only hospitalisations and deaths where COVID-19 was the first coded diagnosis. Higher observed risk of hospitalisation could reflect increased health seeking behaviour (both patient-driven or by a clinician) of those with pre-existing AT use or may indicate that any protective effect from AT materialises only in the most serious cases. The same pattern was observed in AC vs AP and supports the findings from Fröhlich et al that AC may offer more protection against death than AP[9]. For DOACs vs warfarin, no difference was observed between groups for COVID-19 death, but DOACs were associated with marginally reduced odds of COVID-19 hospitalisation. Our analysis did not directly investigate the previously reported observation that vitamin K depletion through warfarin is harmful[15] but, more generally, our findings suggest that it is unlikely that warfarin offers protection against severe COVID-19 outcomes compared with DOACs[11].

Although these protective associations across AT sub-types do not prove causality, they provide further incentive to improve AT coverage for individuals with AF that are already at high risk of stroke. Previous evaluations in the UK have estimated that around 15% of these individuals do not take any AT and around 17% take AP only rather than the recommended AC[3,6]. Our evaluation found around 12% on no AT and around 7% on AP only which suggests national level guidance[16] and primary care incentives such as the Quality and Outcomes Framework [17] continue to have a positive impact. Nonetheless, one in five individuals remains on a sub-optimal medication regime. Shifts from warfarin to DOACs observed in this study and others[18] were recommended by COVID-19 guidance[19] and demonstrate the potential impact of rapidly disseminated medications policy using population scale EHR data.

Identifying which factors are associated with AT use is key to further lowering the proportion of individuals on sub-optimal medication. NSAIDs displayed the strongest association with no AT use and likely reflects the association between NSAIDs and increased risk of major bleeding in individuals with AF[20]. For comorbidities, liver disease had the strongest association with no AT use, which is also supported by clinical evidence[21]. However, recent evidence suggests[22,23] more personalised risk calculations for bleeding and stroke may enable more individuals with liver disease to benefit from AT. History of falls was the comorbidity with the second strongest association with no AT use suggesting it remains a key factor in AT medicating decisions and may be overweighted as a proxy for bleeding risk[7,24,25]. In the UK, NICE guidance was recently updated[4] to explicitly address this issue and it will be important to track the impact of this in future evaluations. On demographics, lower odds of AT use was observed in women but this is likely influenced by using NICE’s primary threshold for the CHA_2_DS_2_-VASc score of 2 for both sexes. The CHA_2_DS_2_-VASc score allocates 1 point to females and 0 for males resulting in a larger proportion of comparatively health females (e.g. 12% and 25% of females in cohort have vascular disease or diabetes vs 21% and 33% respectively in males). However, demographic differences in AT use across ethnicity and socio-economic status mirror systematic healthcare inequalities that have been reported previously[26,27]. Targeted outreach to these groups will be key to improving AT use further.

### Strength and limitations

To our knowledge, this study is the largest scale evaluation of AT use to date. Routinely updated, linked, population-scale EHR datasets provide the statistical power to robustly analyse targeted sub-groups and control for a wide range of potential confounders. The prevalence of individuals with AF and CHA_2_DS_2_-VASc score>=2 in our cohort is similar to that observed in the Quality and Outcomes Framework[17] which provides an external validation for our dataset. All code is opensource and an updated nationwide evaluation can be rapidly created for future time points.

The study does have limitations. Firstly, the reported associations do not demonstrate causality and residual confounding is unlikely to have been fully eliminated. For example, in-hospital treatment regimens were not analysed so differences in COVID-19 outcomes due to additional targeted anticoagulation regimens[28] or other medications such as dexamethasone[29] cannot be accounted for in our analysis. Whilst we attempted to mitigate confounding through careful cohort selection, covariates and propensity score adjustment, our study design does not control for all potential factors associated with the initiation of AT use which may influence COVID-19 outcomes. Lastly, exposure to AT medication was defined as one or more dispensed prescriptions (recorded in NHS BSA Dispensed Medicines) in the previous 6 months. Other studies have used different time periods and prescription frequency counts[9,11] and adherence was not measured[30]. We purposefully defined a liberal threshold to support evaluation of AT usage up to May 2021 that may have included unusual buying patterns (e.g. bulk buying) caused by the pandemic. The trade-off is that for the COVID-19 outcome analyses it increases the probability of including a minority of “exposed” individuals who had ceased regular, pre-existing AT medication.

### Conclusions

Pre-existing AT use may offer marginal protection against COVID-19 death, with AC offering more protection than AP. Although this association may not be causal, it provides further incentive to improve AT coverage for eligible individuals with AF.

## Data Availability

Data are available for bona fide researchers accessible within the NHS Digital trusted Research Environment for England. Contact for information on how to join the CVD-COVID-UK consortium for access.

https://alexhandy1.shinyapps.io/at-evaluation-results/

https://github.com/BHFDSC/CCU020

## ACKNOWLEDGMENTS

This work is carried out with the support of the BHF Data Science Centre led by HDR UK (BHF Grant no. SP/19/3/34678). This work uses data provided by patients and collected by the NHS as part of their care and support. We would also like to acknowledge all data providers who make anonymised data available for research.

The views expressed are those of the authors and not necessarily those of the organisations listed. The funders of this work played no role in the collection, analysis, or interpretation of data; in the writing of the report; or in the decision to submit the article for publication.

## SUPPLEMENTARY MATERIAL

### SUPPLEMENTARY TABLES

**Supplementary Table 1.** study population demographic characteristics for individuals with COVID-19 event by antithrombotic medication category. Percentages should be interpreted vertically for all variables e.g. proportion within category for variable, except for the first row showing percentage of individuals across AT medication categories.

|  | **Total** | **Any AT** | **AC only** | **AP only** | **AC and AP** | **No AT** |
| --- | --- | --- | --- | --- | --- | --- |
| Individuals | 77364<br>(100%) | 67087<br>(86.7%) | 54756<br>(70.8%) | 6743<br>(8.7%) | 5588<br>(7.2%) | 10277<br>(13.3%) |
| Age (mean years, +/- sd) | 81 (+/-<br>10.1) | 81 (+/-<br>9.8) | 81 (+/-<br>9.7) | 82 (+/-<br>10.6) | 79 (+/-<br>9.7) | 81 (+/-<br>12) |
| 65-74 | 12928<br>(16.7%) | 11331<br>(16.9%) | 9123<br>(16.7%) | 1014<br>(15%) | 1194<br>(21.4%) | 1597<br>(15.5%) |
| >=75 | 59369<br>(76.7%) | 51579<br>(76.9%) | 42348<br>(77.3%) | 5289<br>(78.4%) | 3942<br>(70.5%) | 7790<br>(75.8%) |
| Female | 37227<br>(48.1%) | 31498<br>(47%) | 26569<br>(48.5%) | 2992<br>(44.4%) | 1937<br>(34.7%) | 5729<br>(55.7%) |
| Ethnicity |  |  |  |  |  |  |
| White | 72745<br>(94%) | 63132<br>(94.1%) | 51899<br>(94.8%) | 6162<br>(91.4%) | 5071<br>(90.7%) | 9613<br>(93.5%) |
| Asian or Asian British | 2666<br>(3.4%) | 2311<br>(3.4%) | 1578<br>(2.9%) | 389<br>(5.8%) | 344<br>(6.2%) | 355<br>(3.5%) |
| Black or Black British | 1010<br>(1.3%) | 844<br>(1.3%) | 645<br>(1.2%) | 118<br>(1.7%) | 81<br>(1.4%) | 166<br>(1.6%) |
| Mixed | 281<br>(0.4%) | 238<br>(0.4%) | 187<br>(0.3%) | 24<br>(0.4%) | 27<br>(0.5%) | 43 (0.4%) |
| Other Ethnic Groups | 662<br>(0.9%) | 562<br>(0.8%) | 447<br>(0.8%) | 50<br>(0.7%) | 65<br>(1.2%) | 100 (1%) |
| Geographical locations |  |  |  |  |  |  |
| South East | 11387<br>(14.7%) | 9725<br>(14.5%) | 8012<br>(14.6%) | 913<br>(13.5%) | 800<br>(14.3%) | 1662<br>(16.2%) |
| North West | 12691<br>(16.4%) | 11153<br>(16.6%) | 9024<br>(16.5%) | 1113<br>(16.5%) | 1016<br>(18.2%) | 1538<br>(15%) |
| East of England | 7095<br>(9.2%) | 6163<br>(9.2%) | 5047<br>(9.2%) | 591<br>(8.8%) | 525<br>(9.4%) | 932<br>(9.1%) |
| South West | 4185<br>(5.4%) | 3577<br>(5.3%) | 2911<br>(5.3%) | 367<br>(5.4%) | 299<br>(5.4%) | 608<br>(5.9%) |
| Yorkshire and The Humber | 8809<br>(11.4%) | 7639<br>(11.4%) | 6230<br>(11.4%) | 878<br>(13%) | 531<br>(9.5%) | 1170<br>(11.4%) |
| West Midlands | 13273<br>(17.2%) | 11492<br>(17.1%) | 9423<br>(17.2%) | 1129<br>(16.7%) | 940<br>(16.8%) | 1781<br>(17.3%) |
| East Midlands | 7279<br>(9.4%) | 6439<br>(9.6%) | 5376<br>(9.8%) | 603<br>(8.9%) | 460<br>(8.2%) | 840<br>(8.2%) |
| London | 8806<br>(11.4%) | 7528<br>(11.2%) | 5929<br>(10.8%) | 802<br>(11.9%) | 797<br>(14.3%) | 1278<br>(12.4%) |
| North East | 3839<br>(5%) | 3371<br>(5%) | 2804<br>(5.1%) | 347<br>(5.1%) | 220<br>(3.9%) | 468<br>(4.6%) |
| IMD deciles |  |  |  |  |  |  |
| 1 (most deprived) | 8396<br>(10.9%) | 7331<br>(10.9%) | 5843<br>(10.7%) | 832<br>(12.3%) | 656<br>(11.7%) | 1065<br>(10.4%) |
| 10 (least deprived) | 6306<br>(8.2%) | 5449<br>(8.1%) | 4574<br>(8.4%) | 480<br>(7.1%) | 395<br>(7.1%) | 857<br>(8.3%) |

**Supplementary Table 2.**
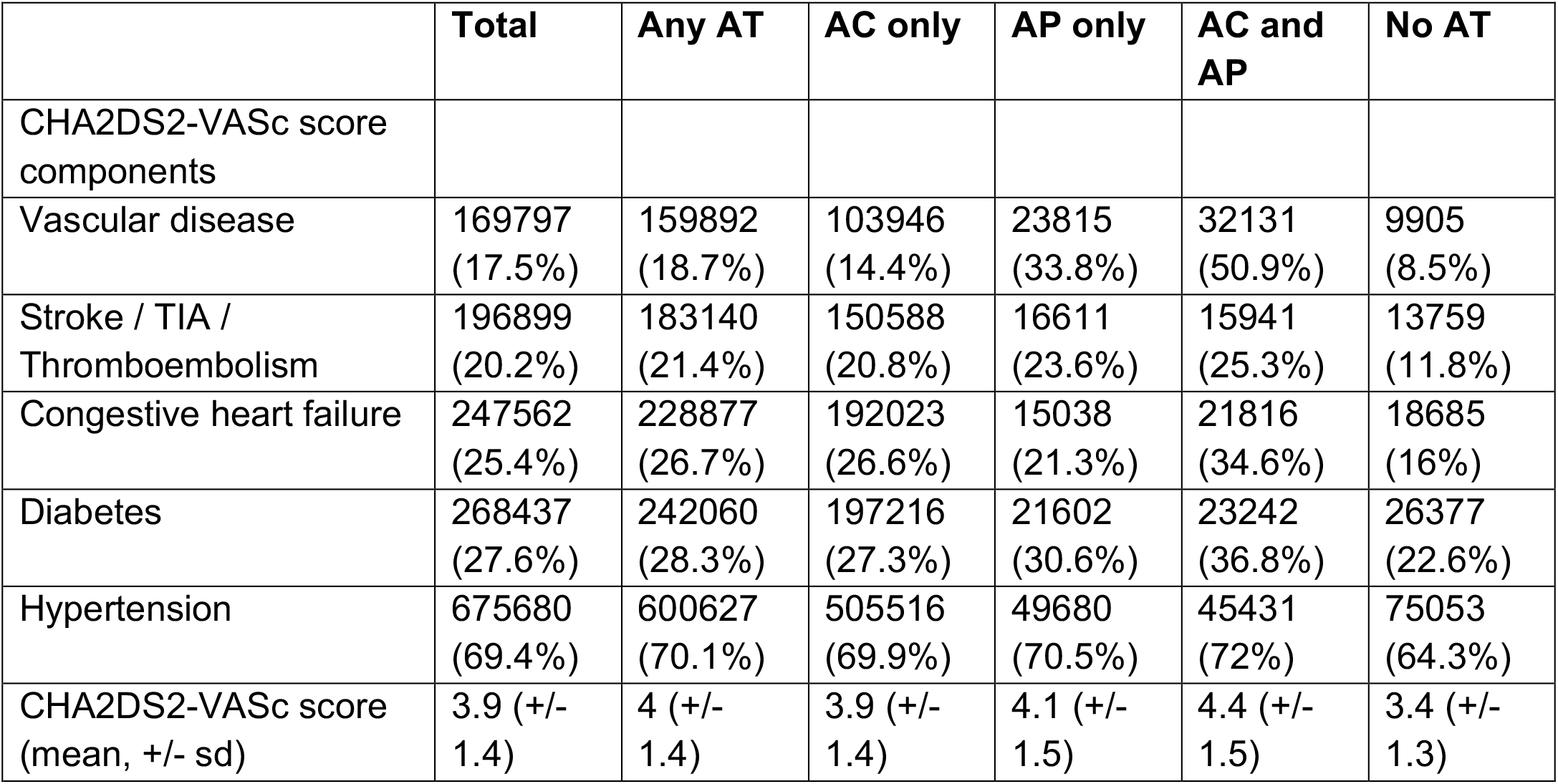
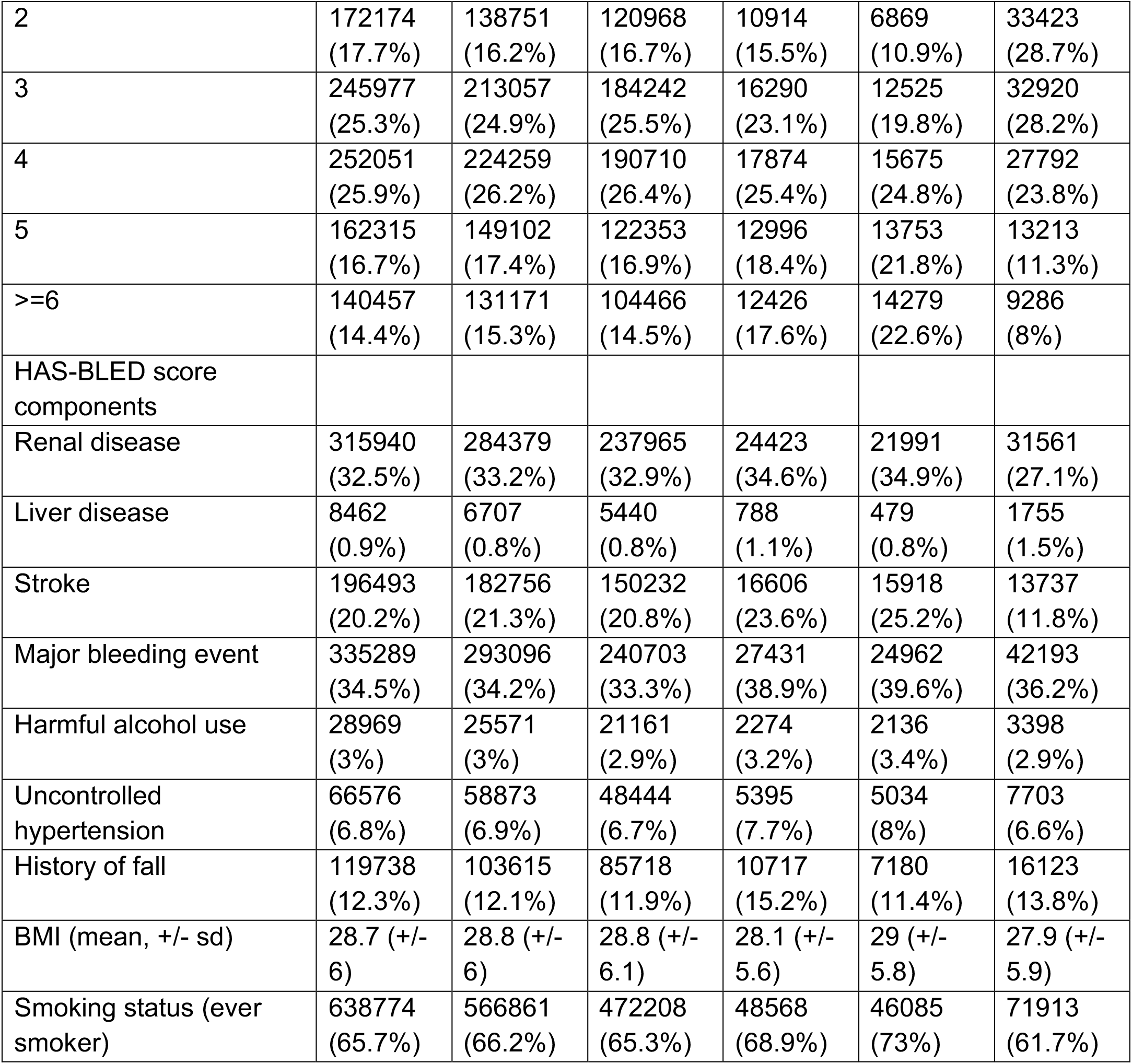
study population comorbidities that increase stroke and bleeding risk for individuals with COVID-19 event by antithrombotic medication category. Percentages should be interpreted vertically for all variables e.g. proportion within category for variable

**Supplementary Table 3.** study population characteristics for COVID-19 outcomes and other medications for individuals with COVID-19 event by antithrombotic medication category. Percentages should be interpreted vertically for all variables e.g. proportion within category for variable

|  | Total | Any AT | AC only | AP only | AC and AP | No AT |
| --- | --- | --- | --- | --- | --- | --- |
| COVID-19 outcomes |  |  |  |  |  |  |
| COVID-19 event | 77364<br>(8%) | 67087<br>(7.8%) | 54756<br>(7.6%) | 6743<br>(9.6%) | 5588<br>(8.9%) | 10277<br>(8.8%) |
| COVID-19 hospitalisation | 37418<br>(3.8%) | 33150<br>(3.9%) | 26887<br>(3.7%) | 3201<br>(4.5%) | 3062<br>(4.9%) | 4268<br>(3.7%) |
| COVID-19 hospitalisation (primary diagnosis) | 27011<br>(2.8%) | 23919<br>(2.8%) | 19375<br>(2.7%) | 2319<br>(3.3%) | 2225<br>(3.5%) | 3092<br>(2.7%) |
| COVID-19 death | 21116<br>(2.2%) | 18173<br>(2.1%) | 14553<br>(2%) | 2055<br>(2.9%) | 1565<br>(2.5%) | 2943<br>(2.5%) |
| COVID-19 death (primary diagnosis) | 15297<br>(1.6%) | 13158<br>(1.5%) | 10522<br>(1.5%) | 1508<br>(2.1%) | 1128<br>(1.8%) | 2139<br>(1.8%) |
| Other medications |  |  |  |  |  |  |
| Antihypertensives | 540681<br>(55.6%) | 498116<br>(58.2%) | 412078<br>(57%) | 40377<br>(57.3%) | 45661<br>(72.4%) | 42565<br>(36.5%) |
| Lipid regulating drugs | 589570<br>(60.6%) | 547522<br>(63.9%) | 441737<br>(61.1%) | 51120<br>(72.5%) | 54665<br>(86.6%) | 42048<br>(36.1%) |
| Proton pump inhibitors | 409430<br>(42.1%) | 369462<br>(43.1%) | 286984<br>(39.7%) | 39181<br>(55.6%) | 43297<br>(68.6%) | 39968<br>(34.3%) |
| NSAIDS | 19448<br>(2%) | 14608<br>(1.7%) | 11101<br>(1.5%) | 2317<br>(3.3%) | 1190<br>(1.9%) | 4840<br>(4.1%) |
| Corticosteroids | 80347<br>(8.3%) | 71706<br>(8.4%) | 59511<br>(8.2%) | 5929<br>(8.4%) | 6266<br>(9.9%) | 8641<br>(7.4%) |
| Other immunosuppressants | 13216<br>(1.4%) | 11690<br>(1.4%) | 9498<br>(1.3%) | 1152<br>(1.6%) | 1040<br>(1.6%) | 1526<br>(1.3%) |
| COVID-19 vaccine prior to COVID-19 event | 9463<br>(1%) | 8248<br>(1%) | 6799<br>(0.9%) | 824<br>(1.2%) | 625 (1%) | 1215<br>(1%) |

### SUPPLEMENTARY FIGURES

**Supplementary Figure 1:**
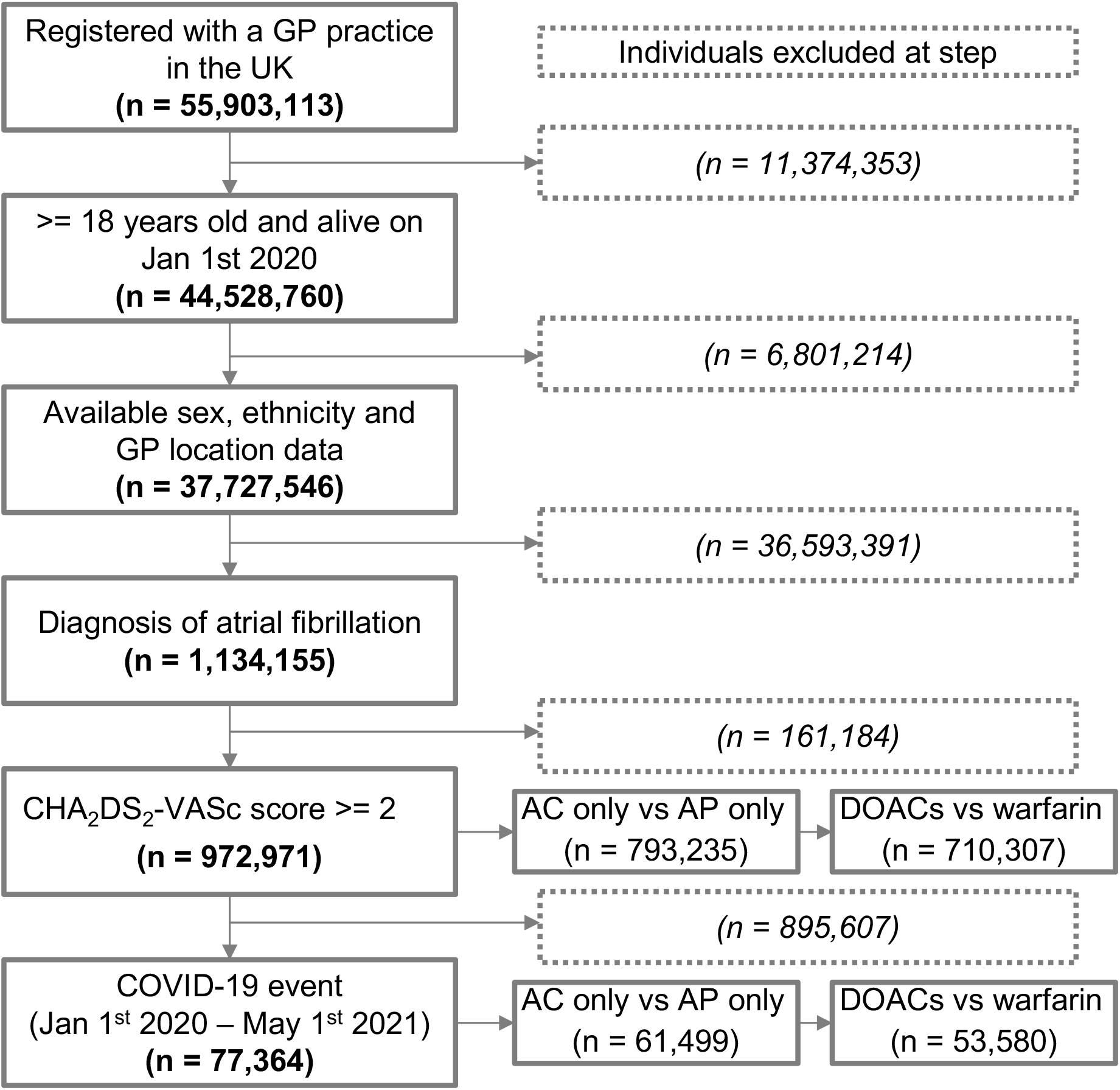
Cohort inclusion flowchart showing the number of individuals excluded at each step and the study population sizes for each question.

**Supplementary Figure 2:**
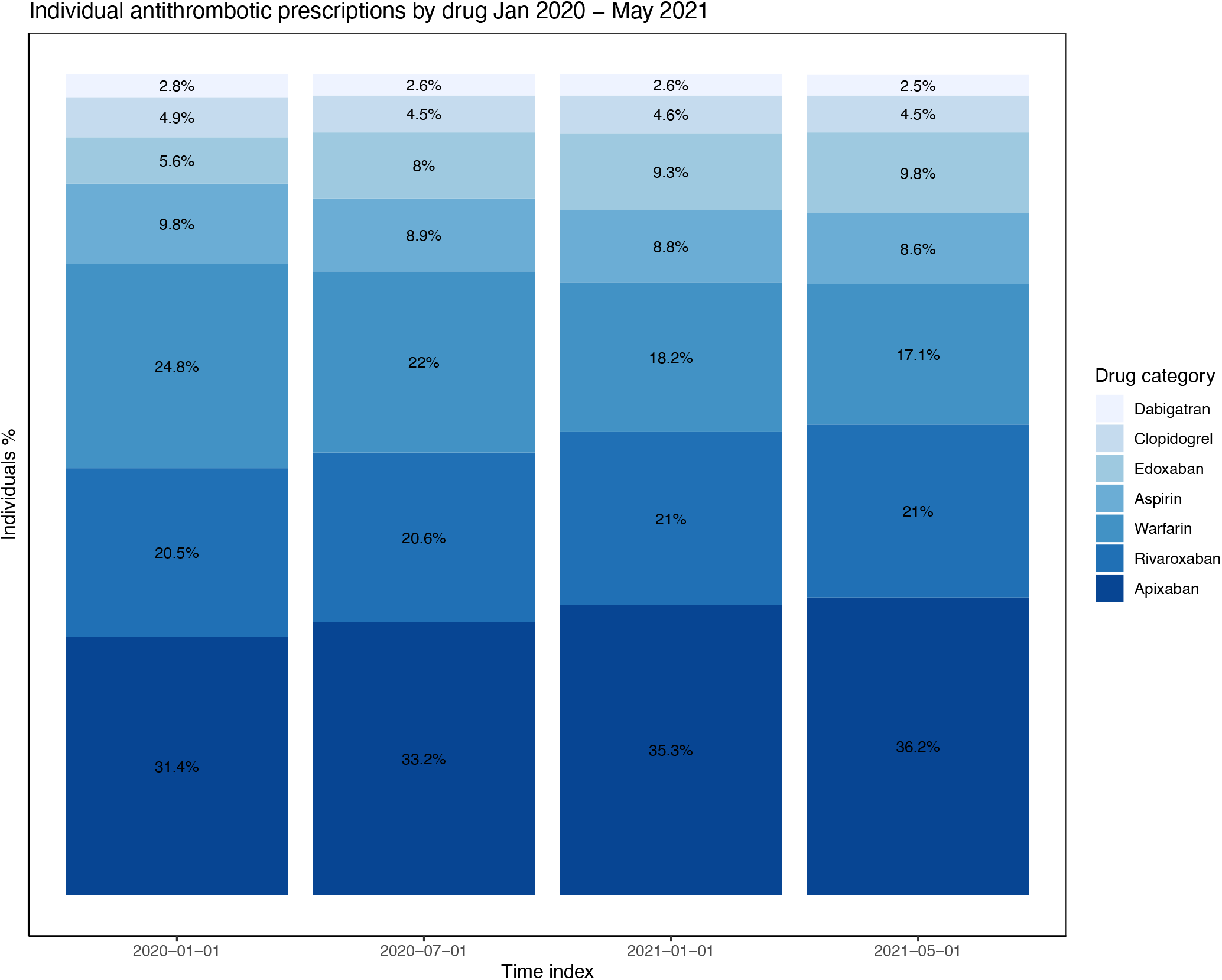
Individual antithrombotic prescriptions by drug January 2020 – May 2021. Ordered by proportion of prescriptions with non-mutually exclusive categories e.g. an individual may have prescriptions for multiple drugs (warfarin and aspirin). Excludes drugs with <1% of prescriptions (ticagrelor, dipyridamole, prasugrel).

**Supplementary Figure 3:**
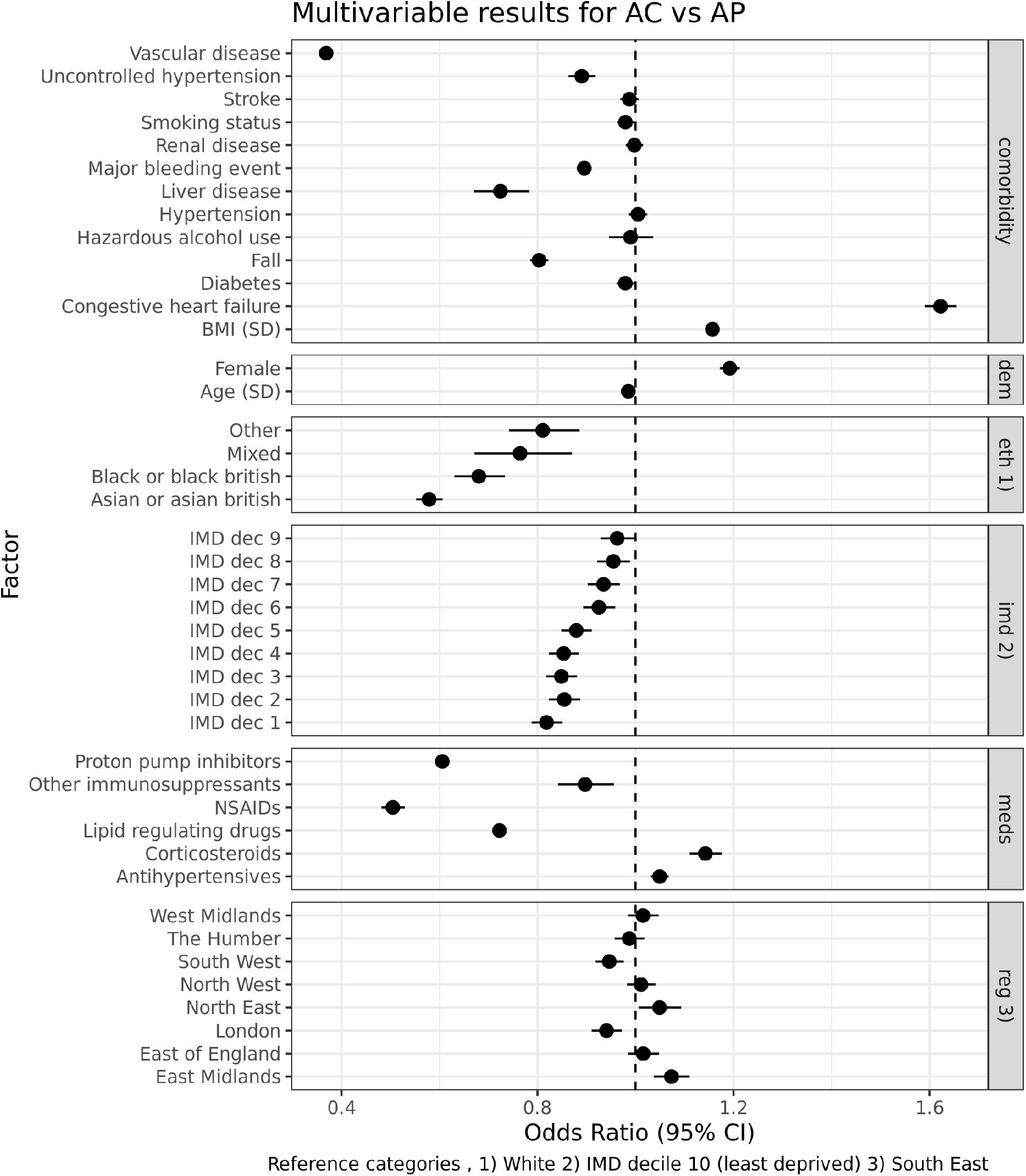
Factors associated with AC vs AP (January 1^st^ 2020) using multivariable logistic regression.

**Supplementary Figure 4:**
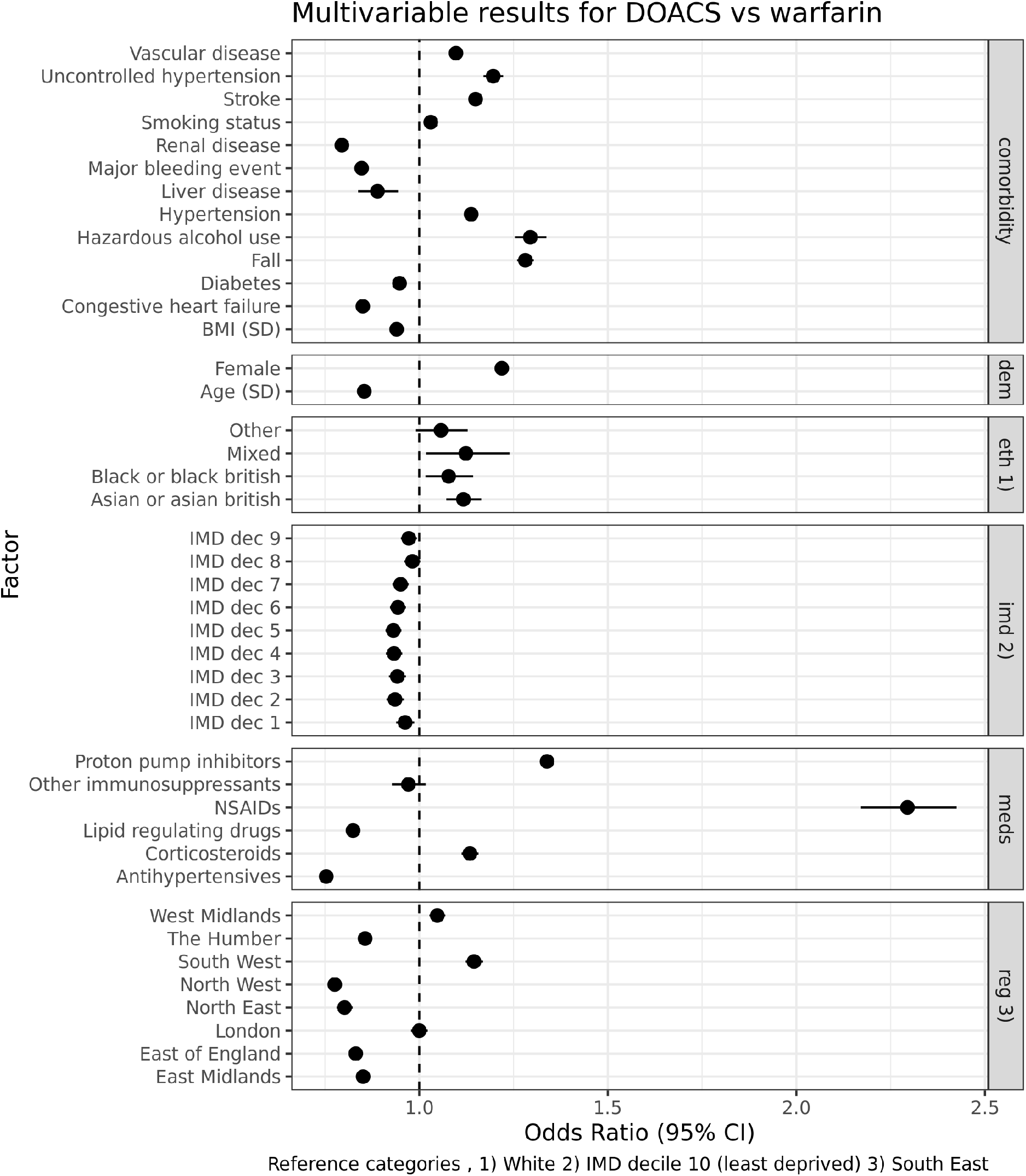
Factors associated with DOACs vs warfarin (January 1^st^ 2020) using multivariable logistic regression.

**Supplementary Figure 5:**
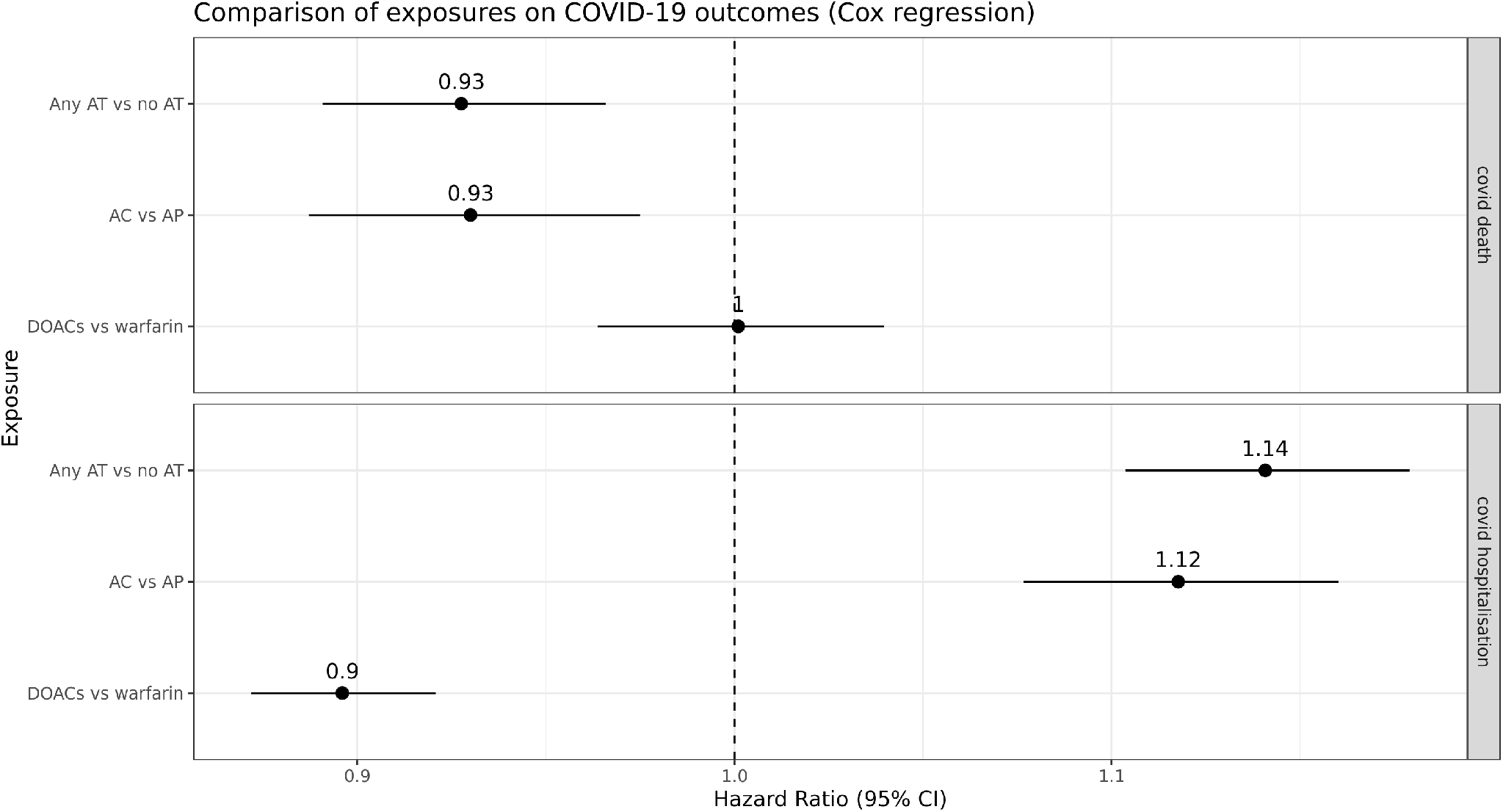
Comparison of AT medication exposures on COVID-19 outcomes (follow up to May 1^st^ 2021) using Cox regression.

**Supplementary Figure 6:**
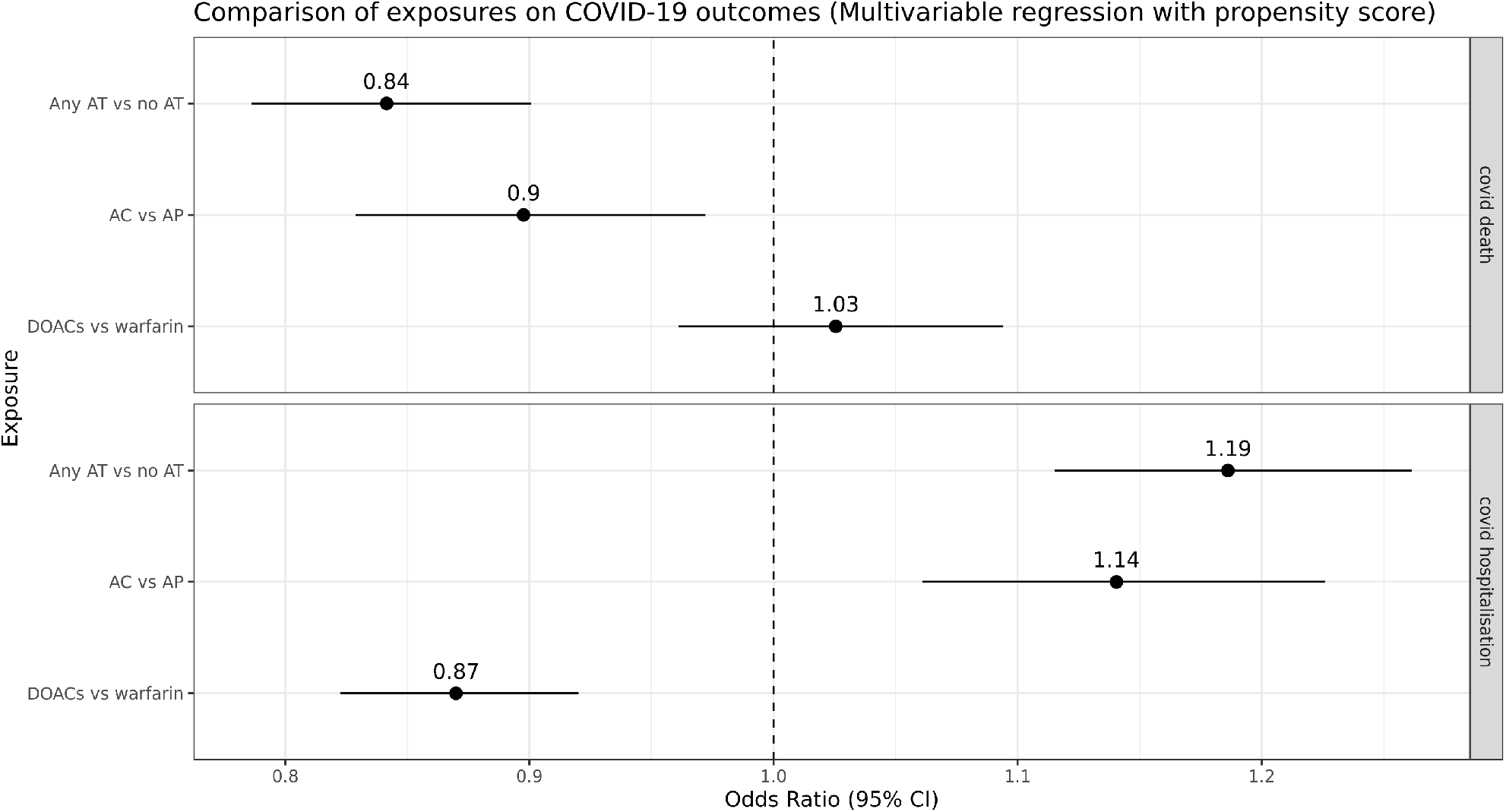
Comparison of AT medication exposures on COVID-19 outcomes (follow up to December 1^st^ 2020) using propensity score adjusted multivariable logistic regression.

**Supplementary Figure 7:**
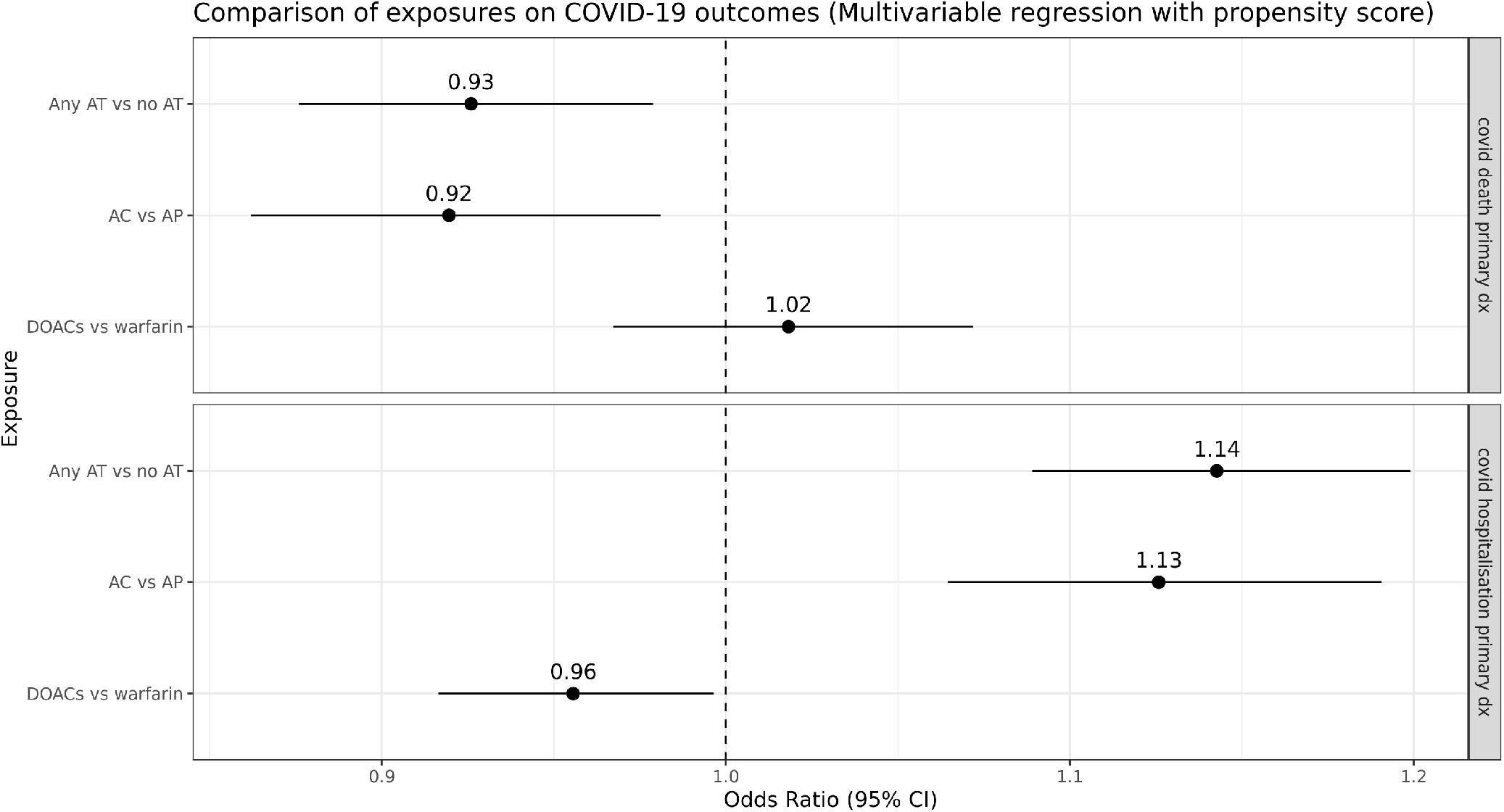
**Comparison of AT medication exposures on COVID-19 hospitalisation and death defined exclusively as the primary recorded diagnosis (follow up to May 1** ^**st**^ **2021) using propensity score adjusted multivariable logistic regression**.

